# Assessing the impact of a gender-neutral approach to HPV vaccination on vaccination coverage for nine-year-old girls in Cameroon: a retrospective, cross-sectional study

**DOI:** 10.64898/2026.04.09.26350560

**Authors:** Bridget C. Griffith, Shariffatou Iliassu, Clarence Mbanga, Budzi Michael Ngenge, Sonali Patel, Justin C. Graves, Navpreet Singh, Shalom Ndoula, Andreas Ateke Njoh, Efouba Gisele, Shadrack Mngemane, Tosin Ajayi, Laure Anais Zultak, Yauba Saidu

**Affiliations:** Analytics and Implementation Research Team, Clinton Health Access Initiative, 383 Dorchester Avenue, Suite 400, Boston, MA 02127, USA; Clinton Health Access Initiative, HS Jean Paul II Boulevard, Tsinga Sous-Prefecture, Yaoundé, Cameroon; Global Vaccines Delivery Team, Clinton Health Access Initiative, 383 Dorchester Avenue, Suite 400, Boston, MA 02127, USA; Expanded Program on Immunization, P.O. Box 2084, Messa, Yaoundé, Cameroon; School of Global Health and Bioethics, Euclid University, Bangui, Central African Republic; Institute for Global Health, University of Siena, Via Valdimontone, 1, 53100 Siena (SI), Italy; College of Health and Health Professions, University of Florida, 1225 Center Drive, Gainesville, FL, 32611 USA

**Keywords:** Gender-Neutral Vaccination, Human Papilloma Virus, HPV Vaccination, Cameroon

## Abstract

Cameroon introduced Human papilloma virus vaccine (HPVV) into the routine immunization schedule in October 2020. By the end of 2022, coverage remained low. To increase coverage, Cameroon switched to a country-wide, gender-neutral vaccination (GNV) approach in 2023, coupled with a revamped delivery strategy consisting of Community Dialogues (CDs) and Periodic Intensification of Routine Immunization (PIRIs) activities in selected health districts (HDs). We assessed the impact of these programmatic changes, notably the GNV approach, on HPVV coverage. This retrospective, cross-sectional study measured the effect of GNV and CDs + PIRIs on HPVV coverage among 9-year-old girls in Cameroon (2022-2023). Data on HPVV coverage from all 203 HDs were extracted from DHIS2, and coverage was calculated at the HD level, based on the estimated population eligible of 9-year-old girls. Descriptive statistics and multiple regression models were employed to assess the impact of GNV on vaccination coverage while adjusting for CDs + PIRIs and urban/rural status. In 2023, of the 203 HDs, 115 (56.7%) conducted GNV only, 74 (36.5%) implemented GNV & CDs + PIRIs, and 75.9% (154) were classified as rural. Among age-eligible girls, there was an overall increase in HPV vaccination coverage, with coverage rising 39.2 percentage points from 2022 to 2023. Following multiple linear regression, there was a significant increase in HPVV coverage in HDs with GNV & CDs + PIRIs compared to those with no GNV and no CDs + PIRIs (β:55.5%, 95%CI: 38.7, 72.3, *p*=0.000). Furthermore, there was a significant increase in HPVV coverage in HDs with GNV only compared to those with no GNV or no CDs + PIRIs (β:28.7%, 95%CI: 12.5, 45.0 *p*=0.001).

Overall, the GNV approach increased HPVV coverage for girls significantly, particularly when implemented alongside CDs + PIRIs.

## Introduction

Human papillomavirus (HPV) is the most prevalent sexually transmitted infection worldwide, with approximately 80% of adults acquiring the virus during their lifetime [1, 2]. The disease is caused by one or more of the nearly 200 existing HPV serotypes. At least, 12 of these serotypes are known to be oncogenic, causing several types of cancers, including cervical, anal, oropharyngeal, vulvar, vaginal, and penile cancers, [3, 4]. The virus is responsible for nearly 99% of cervical cancer cases, which is the fourth most common cancer among women globally [3, 5]. In 2022, over 660,000 new cervical cancer cases and 350,000 related deaths were recorded, with more than 90% of these deaths occurring in low-and middle-income countries (LMICs), particularly in sub-Saharan Africa [3].

The HPV vaccine (HPVV) prevents HPV infection and can lower the risk of developing HPV related cancers [6]. The HPVV first became available for females in the United Staes in 2006 [7]. As of December 2025, 156 countries and territories have the HPVV on their national routine immunization schedule [8–10]. Globally, HPV vaccination coverage increased from 13% in 2019 to 21% in 2024 (first dose by age 15, females) and from 11% in 2019 to 18% in 2024 (last dose by age 15, females) [10, 11].

Despite the availability of safe and effective HPV vaccines, coverage remains low in Sub-Saharan Africa, where the burden of HPV-related disease is highest [12]. This persistently low coverage has been attributed to multiple barriers to vaccine uptake, including supply chain limitations, misinformation, vaccine hesitancy, and sociocultural factors, which are particularly pronounced in LMICs and sub-Saharan Africa [13–16]. Innovative strategies are needed to overcome these barriers and expand vaccine coverage.

In Cameroon, cervical cancer ranks as the second most common cancer among women, with approximately 2,770 new cases and 1,770 deaths annually [17]. To combat this high burden, Cameroon introduced a two-dose schedule of the HPVV into the routine immunization program in October 2020, targeting nine-year-old girls. However, coverage remained critically low, with coverage for the first and second dose estimated at 20.3% and 5.3% respectively by December 2022, far below the nationally defined target coverage of 40% at the time of introduction. Published reports suggest that this low uptake was, in part, driven by, insufficient stakeholder engagement, and spillover misinformation from COVID-19 vaccination, multiple dose schedules, misconceptions about the vaccine, including the fact that it is tool designed to sterilized young girls[18, 19].

To address these challenges with HPVV uptake, Cameroon’s National Immunization Technical Advisory Group (NITAG) recommended transitioning to a one-dose schedule in November 2022, following evidence from global studies demonstrating its comparable efficacy to a two-dose regimen [20]. Additionally, Cameroon’s NITAG advised implementing a gender-neutral vaccination (GNV) strategy to vaccinate boys alongside girls, making Cameroon the first LMIC to introduce GNV for HPVV. This change is based on the recognition of the potential for HPV transmission by men and the low female vaccination coverage [21, 22]. This decision was backed by evidence from high-income settings, which suggest that vaccinating boys amplifies the public health impact of HPV vaccines and addresses sociocultural barriers tied to girl-only vaccination strategies [23].

The GNV strategy and one-dose schedule were rolled out nationwide in January 2023. To support and complement this transition, the Expanded Program on Immunization (EPI) implemented targeted interventions, including Periodic Intensification of Routine Immunization (PIRI) campaigns and community dialogues (CDs) in selected health districts (HDs), to address vaccine hesitancy and improve uptake among previously missed populations. These interventions aimed to expand vaccination coverage by engaging community leaders, addressing misconceptions, and integrating facility- and community-based vaccination efforts.

Given the novelty of the GNV strategy in Cameroon, and more broadly in LMICs, and the reliance on interventions targeting community engagement, there was a need to assess the effectiveness of these approaches in improving HPVV coverage. The study aimed to describe the distribution of GNV and CDs/PIRIs in Cameroon in 2023, assess the change in HPVV coverage among eligible girls in Cameroon pre (2022) and post (2023) the implementation of these approaches, and evaluate the impact of the approaches (GNV and GNV + PIRIS and CDs) on the change in HPVV coverage. To achieve these aims, we conducted a retrospective analysis of administrative and programmatic data. This study provides insights into the implementation outcomes and effectiveness of these strategies, contributing to the evidence required to inform sustainable vaccination policies in LMICs.

## Methods

### Study design, setting, and participants

We conducted a retrospective, cross-sectional analysis of national administrative HPV vaccine coverage data in Cameroon to assess the effect of GNV and CDs followed by PIRIs on HPV vaccination coverage among age-eligible girls in Cameroon. Quantitative data, spanning a 24-month period (January 2022 to December 2023), were extracted from the Cameroon District Health Information System (DHIS2) to evaluate changes in HPV vaccination coverage before and after the implementation of the GNV strategy in January 2023, alongside the use of CDs and PIRIs for HPV vaccination.

The study included data from all 203 HDs in all ten regions of Cameroon, because the implementation of HPVV-related interventions (CDs, PIRIS, and GNV) in 2023 was country-wide. HDs are distributed across the ten regions of Cameroon, with the highest number of HDs located in the Extreme-North region of the country. Across Cameroon, CDs and PIRIs were either organized by the national EPI or sub nationally by HD-level leadership. The designation of urban and rural HDs is as defined and designated by the Cameroon National Institute of Statistics.

### Ethical considerations

Prior to protocol development, study PIs held validation meetings with national EPI to align on the study scope, study aims, and intended data use. The study protocol was reviewed and approved by the Clinton Health access Initiative’s internal Scientific and Ethical Review Committee (SERC). It was also approved by Cameroon’s National Ethics Committee for Human Health Research (CNERSH) (application number 2024/06/168666/CE/CNERSH/SP). The data was accessed from the Cameroon DHIS2 on the 30th of June 2024, and the authors had no access to any information that could identify individuals during or after the data collection.

### Definitions

Vaccination for HPV was defined as having received one dose of HPV vaccination within the calendar year (HPV1).

The population of children who are eligible for HPV vaccination is based on the estimated number of 9-year-old girls and boys within each HD. Target populations (number of eligible girls, number of eligible boys) as calculated by the EPI, were estimated based on the number of 9-year-old girls and boys in the Cameroon 2005 census data, adjusted for population growth over time.

To confirm the presence of CDs, followed by PIRIs in each HD, we conducted a review of all relevant documents (including NITAG reports, EPI monthly, and EPI annual reports) pertaining to GNV CDs and PIRIs conducted. To confirm if sub-national CDs + PIRIs occurred, regional immunization coordinators were consulted to identify any HDs where HD-level HPV-related PIRI initiatives were conducted beyond the nationally implemented ones. In addition, we held calls with all EPI regional coordinators to check the implementation of sub-national (HD-level) CDs and PIRIs. The occurrence of CDs and PIRIs is referred to as “CDs + PIRIs” in the analysis.

The implementation of GNV for HPV was initiated through a nationwide policy change, but in order to determine whether GNV occurred in a HD, the number of eligible boys vaccinated was collected. If zero eligible boys were vaccinated for HPV in 2023, the HD was categorized as having “no GNV” occur.

### Data collection, management, and validation

Data were extracted from the Cameroon DHIS2 immunization portal for all 203 HDs with permission from the EPI Permanent Secretary. The data consisted of two years (2022-2023) anonymized administrative records, aggregated by Health Area, with no linkage to individual private information, ensuring confidentiality. The dataset included HPVV nationwide coverage for age-eligible girls (years 2022-2023) and boys (year 2023) by Health Area. To maintain data security, all files were stored on a password-protected server, accessible only to authorized personnel using individual logins and role-based permissions. Before commencing data extraction procedures, approvals were obtained from all relevant administrative bodies at national and sub-national levels. Upon completing the analysis, data files were deleted to ensure confidentiality and compliance with ethical standards.

We conducted data cleaning to identify and recode missing values, check for outliers and potential data entry errors, and address inconsistencies in formatting. Data cleaning, analysis, and visualization was conducted with Stata (Version 16) and Excel (Microsoft 365).

### Statistical analysis

HPVV coverage amongst eligible girls was calculated as the number of eligible girls vaccinated with one dose of the HPVV in each HD divided by the estimated number of 9-year-old girls in each HD. The same approach was used to calculate HPVV coverage amongst eligible boys.

Descriptive characteristics of HDs and interventions were summarized using frequencies and percentages. Vaccination totals and proportions were computed by summing the number of targeted and vaccinated girls annually, stratified by region, HD, and rural versus urban status. Due to variations in CDs and PIRIs implementation at the health area level, the unit of analysis was aggregated to the HD.

To assess the relationship between GNV, CDs and PIRIs and change in HPVV coverage among girls, we conducted two multiple linear regression models with change in HPVV coverage in girls from 2022 to 2023 as the continuous outcome. Variables were selected for inclusion in the model *a priori,* and model fit (R^2^) was considered in selection of final models. We present adjusted and unadjusted model outputs.

In the first model, the dependent variable was the change in HPVV coverage in girls from 2022 to 2023, and the independent variables were intervention (no GNV or CDs + PIRIs (ref); GNV+CDs + PIRIs; GNV only) and urban/rural status(urban(ref); rural).

In the second model, the dependent variable was the change in HPVV coverage in girls from 2022 to 2023, and the independent variables were GNV (no (ref); yes), CDs + PIRIs (no (ref); yes), and urban/rural status(urban(ref); rural).

We conducted sensitivity analyses to determine the effect of vaccine coverage values above 100% on the model results and interpretation. To do this, we ran the model first with coverage values over 100%, then with the coverage values over 100% set to missing, and third with the coverage values capped at 100%. The outputs of the models in which vaccine coverage values were capped at 100% are presented in this manuscript.

## Results

### HPV Vaccination Coverage in Cameroon 2022-2023

In 2022, of the 415,296 nine-year old girls targeted for vaccination with HPVV, 80,168 (19.3%) were vaccinated with one dose of HPVV. In 2023, 249,594 (58.5%) out of 426,429 girls targeted were vaccinated with one dose of HPVV. In 2023, 112,606 (26.9%) of the 417,985 boys who were eligible for HPVV were vaccinated (Table 1).

**Table 1:** Number and proportion vaccinated for HPVV.

|  | (Number vaccinated/Number eligible); HPV coverage |  | HPV coverage percentage point difference between 2022 and 2023, 95% CI |
| --- | --- | --- | --- |
| Intervention (number of health districts) | Year |  |  |
| Overall (n=203) | 2022 | 2023 |  |
| Girls | (80,168/425,296); 19.3% | (249,594/426,429); 58.5% | 39.2% (39.0, 39.4) |
| Boys | - | (112,606/417,985); 26.9% | - |
| GNV (n=115) | 2022 | 2023 |  |
| Girls | (32,445/238,654); 13.6% | (80,829/245,442); 32.9% | 19.3% (19.1, 19.5) |
| Boys | - | (34,988/240,581); 14.5% | - |
| GNV & CDs + PIRs (n=74) | 2022 | 2023 |  |
| Girls | (47,219/166,953); 28.3% | (168,222/171,168); 98.5% | 70.2% (70.0, 70.4) |
| Boys | - | (77,618/167,778); 46.3% | - |
| No GNV or CDs + PIRs (n=14) | 2022 | 2023 |  |
| Girls | (504/9697); 5.2% | (143/9820); 1.5% | -3.7% (3.2, 4.2) |
| Boys | - | (0/9625); 0% | - |

In the 115 HDs where only GNV was implemented, the HPVV coverage increased from 13.6% in 2022 to 32.9% in 2023, a 19.3 percentage point increase. A larger increase in coverage from 2022 to 2023 was observed among the 74 HDs where GNV was implemented alongside CDs + PIRIs, with an increase in vaccination coverage from 28.3% in 2022 to 98.5% in 2023, a 70.2 percentage point increase (Table 1).

As presented in Table 2, among all HDs, the median proportion of eligible girls vaccinated for HPV in 2022 was 9.6% (IQR: 1.7, 31.0). This median proportion increased to 47.5% (IQR: 7.3, 101.0) in 2023. The greatest increase in coverage between 2022 and 2023 was observed amongst the HDs where both GNV and CDs + PIRIs occurred (68.5%; IQR: 36.6, 126.0). HDs that only implemented GNV also had an increase in coverage of 13.0% (IQR: 1.7, 38.2), while HDs that did not implement GNV or CDs + PIRIs had a minimal change in coverage, which was a decrease in some HDs, between the two years (0.27%; IQR: -1.3, 1.9).

**Table 2:**
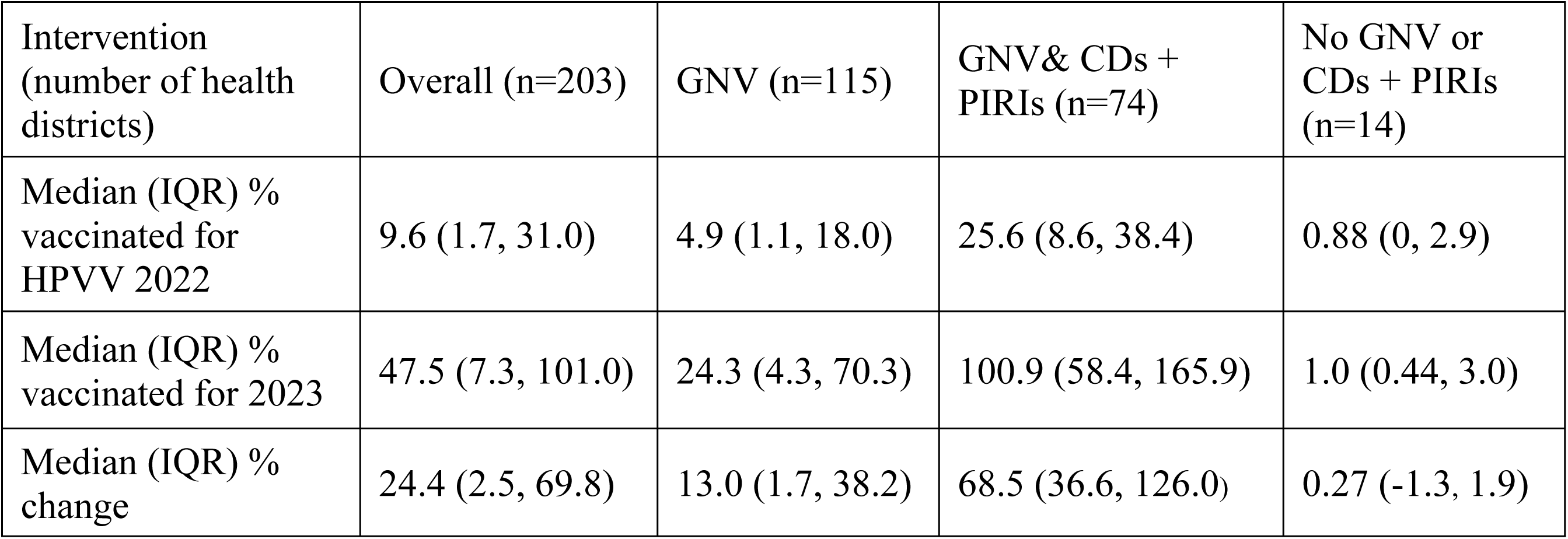
Median HPVV coverage for girls, per GNV intervention.

### Distribution of GNV and CDs + PIRIs

As presented in Table 3, out of the 203 HDs, about half (56.7%; n=115) conducted GNV of HPVV for eligible children in 2023 without CDs + PIRIs. In about one third (36.5%; n= 74) of HDs, GNV was implemented alongside CDs + PIRIs, organized either at the national-level or HD-level. The remaining 14 HDs (6.9%) did not implement GNV of HPVV (no eligible boys were vaccinated) or conduct CDs + PIRIs. Just over one fifth of the HDs (21.2%; n=43) are classified as urban, while the rest are classified as rural (78.8%; n=160).

**Table 3:** Description of health districts, by intervention.

| **Table 3: Description of health districts, by intervention** |  |  |  |  |
| --- | --- | --- | --- | --- |
|  | Number of health districts |  |  |  |
|  | Overall<br>n (column<br>%) | Implemented<br>GNV only | Implemented GNV<br>and CDs + PIRIs | Implemented<br>no GNV or<br>CDs + PIRIs |
| Number of HDs<br>n (row %) | 203 (100) | 115 (56.7) | 74 (36.5) | 14 (6.9) |
| Region<br>n (row %) |  |  |  |  |
| Adamawa | 11 (5.4) | 4 (36.4) | 7 (63.6) | 0 (0.0) |
| Centre | 32 (15.8) | 20(62.5) | 7 (21.9) | 5 (15.6) |
| East | 15 (7.4) | 8 (53.3) | 7 (46.7) | 0 (0.0) |
| Extreme-North | 32 (15.8) | 18 (56.3) | 14 (43.8) | 0 (0.0) |
| Littoral | 24 (11.8) | 13 (54.2) | 7 (29.2) | 4 (16.7) |
| North | 15 (7.4) | 7 (46.7) | 8 (53.3) | 0 (0.0) |
| North-West | 21 (10.3) | 16 (76.2) | 4 (19.1) | 1 (4.8) |
| West | 20 (9.9) | 9 (45.0) | 7 (35.0) | 4 (20.0) |
| South | 12 (5.9) | 7 (58.3) | 5 (41.7) | 0 (0.0) |
| South-West | 21 (10.3) | 13 (61.9) | 8 (38.1) | 0 (0.0) |
| Median proportion<br>(IQR) |  | 55.3 (15.8) | 39.9 (21.0) | 0.0 (15.9) |
| Urbanicity<br>n (row %) |  |  |  |  |
| Rural | 160 (78.8) | 94 (58.8) | 54 (33.8) | 12 (7.5) |
| Urban | 43 (21.2) | 21 (48.8) | 20 (46.5) | 2 (4.7) |

The proportion of HDs within a region that conducted each of the three implementation types (GNV only, GNV & CDs + PIRIs, and no GNV or CDs + PIRIs) varied across all regions, with a median of 55.3% (IQR=15.8) of HDs per region involved in GNV only, 39.9% (IQR=21.0) in GNV & CDs + PIRIs, and 0.0% (IQR=15.9) in no GNV or CDs + PIRIs.

### Effect of GNV and CDs + PIRIs on HPVV coverage

To measure the effect that GNV and GNV & CDs + PIRIs had on the change in HPVV coverage for girls, we conducted a multiple linear regression model, with change in vaccine coverage from 2022 to 2023 as the continuous outcome. We present the outputs of the models (Table 4, 5) in which vaccine coverage values were capped at 100%, because the overall directionality of the associations did not change within the sensitivity analysis and the best model fit was observed when vaccine coverage values were capped at 100%.

**Table 4:** Multiple linear regression of the combined effect of GNV & CDs + PIRIs on change in HPVV coverage output.

| Table 4: Multiple linear regression of the combined effect of GNV & CDs + PIRIs on change in HPV coverage output |  |  |  |  |  |
| --- | --- | --- | --- | --- | --- |
| Dependent: Change in HPV coverage |  |  |  |  |  |
|  | N (%) | Coef. (Unadjusted) | 95% CI; p-value | Coef. (Adjusted) | 95% CI; p-value |
| No GNV or CDs + PIRIs | 14 (6.9) | ref. |  | - |  |
| GNV & CDs + PIRIs | 74 (36.5) | 53.8 | 36.8, 70.8; 0.000 | 55.5 | 38.7, 72.3; 0.000 |
| GNV only | 115 (56.7) | 28.2 | 11.7, 44.7; 0.001 | 28.7 | 12.5, 45.0; 0.001 |
| Rural | 160 (78.8) | ref. |  | ref. |  |
| Urban | 43 (21.2) | 0.07 | -0.03, 0.18; 0.169 | -13.8 | -23.7, -3.8; 0.007 |

In the model assessing the relationship between the presence of GNV & CDs + PIRIs and change in vaccine coverage, HDs with GNV & CDs ++ PIRIs were predicted to have a 55.5 percentage point increase in coverage change (95%CI: 38.7, 72.3), compared to HDs with no GNV or CDs + PIRIs, while controlling for urban/rural status. Furthermore, HDs with GNV only were predicted to have a 28.7 percentage point increase in vaccine coverage (95%CI: 12.5, 45.0), when compared to HDs with no GNV or CDs + PIRIs (Table 4, Figure 1).

**Figure 1:**
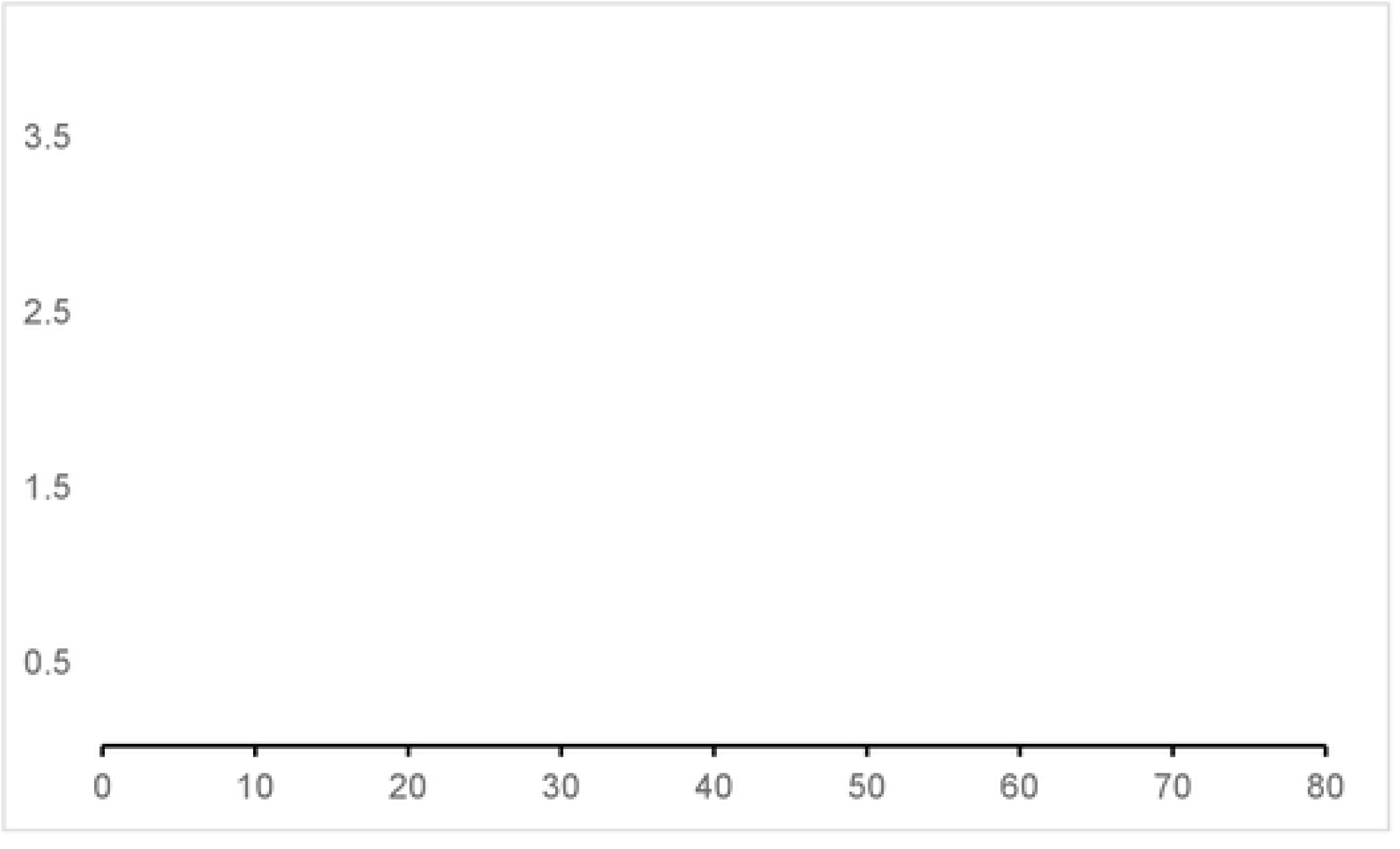
Adjusted model output (coefficients and 95% confidence intervals) from multiple linear regression models. Model output from the models assessing the relationship between the listed independent variables and change in HPVV coverage among girls. *Indicates adjusted model output

We also assessed the effect of CDs + PIRIs and GNV individually on the difference in HPV vaccine coverage in 2022 and 2023. To do this, we ran an additional multiple linear regression model with change in vaccine coverage as the continuous outcome, and GNV (yes/no), CDs + PIRIs (yes/no) as independent variable, in addition to controlling for urban/rural status (urban/rural/NA). HDs with GNV were predicted to have a 28.7 percentage point increase in coverage change (95%CI: 12.5, 45.0), compared to HDs with no GNV. HDs with CDs + PIRIs were predicted to have a 26.8 percentage point increase in vaccine coverage (95%CI: 18.2, 35.4), when compared to districts with no CDs + PIRIs (Table 5, Figure 1).

**Table 5:** Multiple linear regression of the individual effect of GNV and CDs+ PIRIs on change in HPVV coverage output.

| Table 5: Multiple linear regression of the individual effect of GNV and CDs+ PIRIs on change in HPV coverage output |  |  |  |  |  |
| --- | --- | --- | --- | --- | --- |
| Dependent: Change in HPV coverage |  |  |  |  |  |
|  | N (%) | Coef. (Unadjusted) | 95% CI; p-value | Coef. (Adjusted) | 95% CI; p-value |
| No GNV | 14 (6.9) | ref. |  | - | - |
| GNV | 189 (93.1) | 38.2 | 20.8, 55.6; 0.000 | 28.7 | 12.5, 45.0; 0.001 |
| No CDs + PIRIs | 129 (63.6) | ref. |  | - | - |
| CDs + PIRIs | 74 (36.5) | 28.6 | 19.9, 37.4; 0.000 | 26.8 | 18.2, 35.4; 0.000 |
| Rural | 160 (78.8) | ref. | - | ref. | - |
| Urban | 43 (21.2) | 0.07 | -0.03, 0.18; 0.169 | -13.8 | -23.7, -3.8; 0.007 |

## Discussion

We conducted a retrospective analysis of HPVV data from 2022 to 2023 to determine the effect that GNV and PIRIs & CDs had on HPVV coverage for eligible-aged girls in Cameroon. This study highlights the positive impact of the Gender-Neutral Vaccination (GNV) approach on HPV vaccination coverage, suggesting a substantial increase in coverage for girls in 2023.

While GNV increased the proportion of girls vaccinated for HPV, the combination of GNV and CDs + PIRIs led to a significantly larger increase in the proportion vaccinated, based on the outputs of multiple linear regression models. This suggests that CDs + PIRIs play a key role in substantial coverage gains on top of the GNV policy and represent an important component of national and subnational vaccination efforts. Carrying out frequent CDs and PIRIs promotes interaction and engagement with community members and Civil Society Organizations (CSOs), allowing for the handling of issues and misconceptions about vaccination, and ultimately enhancing acceptance and uptake among families[24, 25]. Furthermore, this analysis demonstrates that securing sufficient funding and resources is a critical investment for effective implementation, outreach, and sustained engagement in vaccination initiatives [26]. A previously published study on the GNV strategy for HPVV in Cameroon describes the HPVV implementation process, GNV rollout and CDs + PIRIs timeline in detail [22]. This study suggests that PIRIs are critical in significantly increasing the uptake of HPVV among both boys and girls in Cameroon, using a segmented regression with interrupted time series methodology. In addition, this study estimated HPVV coverage at the regional level, highlighting that the WHO recommend threshold for HPVV coverage of 90% was achieved in some regions as a result of the combined efforts of GNV, CDs, and PIRIs[5, 22]. Our study builds on this work by describing the effect of GNV and CDs + PIRIs on change in HPVV coverage at a national level and demonstrating a method for managing out of range vaccine coverage estimates using sensitivity analysis.

Gender neutral HPV vaccination strategies are identified as a more inclusive approach to disease prevention, including evidence that vaccination of boys/men can reduce the transmission of HPV[27] and reduce the risk of HPV-related cancers in men[28]. This is also motivated by the estimate that 80% HPVV coverage is needed among girls to reduce HPV infection in boys, and current global and regional coverage remains lower than that threshold[6]. This inclusive approach has been highlighted as an effective strategy to comprehensively address HPV-driven disease and improve coverage of HPVV in sub-Saharan Africa[29], but there is minimal evidence for or against the expansion of HPV vaccination to include boys/men specifically to increase HPVV uptake among girls. This is among one the first studies to assess the impact of GNV on HPVV coverage among girls.

There remains a debate about the cost effectiveness of expanding HPV vaccination to boys/men, in which cost effectiveness depends heavily on the vaccine coverage of females [30]. In addition, economic evaluations of the cost effectiveness in LMICs and sub-Saharan Africa are extremely limited. While this study does not directly address costs, it does demonstrate a potential practical benefit to vaccinating boys (an increase in uptake among girls). Future studies of real-world implementation on gender neutral HPVV should include a measure of the change in HPVV uptake among girls. In addition, an increase in the number of years of data will improve the precision of these findings.

This study has several limitations. The observational design lacks randomization, which may have introduced selection bias and limited the ability to establish causal relationships between the GNV approach and vaccination outcomes. In addition, the lack of randomization meant the intervention areas were not balanced, and the places without CDs + PIRIs could possibly be correlated with low coverage or other coverage-influencing factors. The lack of visibility into these factors may cause unaccounted for biases in the analysis and interpretation. In this setting, CDs + PIRIs were always done in tandem, which is why they are grouped together throughout the analysis. Therefore, we were not able to differentiate the effects of CDs or PIRIs individually.

Our study relied on retrospective administrative data from DHIS2 that was not collected for research; it contained some missing and inconsistent values, such as coverage estimates over 100%. We addressed these challenges by changing the unit of analysis from health area to HD and conducting sensitivity analyses to ascertain the effect the nonsensical values had on model outputs. In addition, administrative data reporting the presence of CDs + PIRIs by geographical unit may lead to mis-categorization and bias, as the reality of the presence of these interventions may be different at the individual level.

## Conclusion

This analysis of administrative data demonstrates that the Gender-Neutral Vaccination (GNV) approach proved to be an effective strategy in increasing HPV vaccination coverage in girls, particularly when accompanied with CDs and PIRIs. The implications of this study may be useful for other countries considering a GNV strategy to address low HPVV coverage, especially when low coverage is due to gender-related rumours. Future research should focus on measuring this relationship over longer periods of time and further differentiating between the effects that GNV, CDs, PIRIs, and a combined approach has on HPVV uptake. Overall, investment in research will be crucial for sustaining and expanding the positive impacts of the GNV approach, ensuring that no child is left unprotected against HPV-related cancers.

## Data Availability

The datasets generated and analyzed during the current study are available from the corresponding author on request.

## Acknowledgments

The authors would like to express their sincere appreciation for the invaluable support provided by the members of the Central and Regional EPI teams, as well as the leadership of the respective target HDs. Their facilitation, coordination, and commitment were essential in ensuring the smooth implementation of this study across the selected regions and districts.

## References

1. Chesson HW, Dunne EF, Hariri S, Markowitz LE. The estimated lifetime probability of acquiring human papillomavirus in the United States. Sexually transmitted diseases. 2014;41(11):660–4.

2. Forman D, de Martel C, Lacey CJ, Soerjomataram I, Lortet-Tieulent J, Bruni L, et al. Global burden of human papillomavirus and related diseases. Vaccine. 2012;30 Suppl 5:F12–23. doi: 10.1016/j.vaccine.2012.07.055. PubMed PMID: 23199955.

3. World Health Organization (WHO)/Organisation mondiale de la Santé (OMS). Cervical cancer 2024. Available from: https://www.who.int/news-room/fact-sheets/detail/cervical-cancer.

4. Bouvard V, Baan R, Straif K, Grosse Y, Secretan B, El Ghissassi F, et al. A review of human carcinogens—Part B: biological agents. The lancet oncology. 2009;10(4):321–2.

5. World Health Organization (WHO)/Organisation mondiale de la Santé (OMS). Global strategy to accelerate the elimination of cervical cancer as a public health problem World Health Organization (WHO), editor. Geneva2020.

6. World Health Organization (WHO)/Organisation mondiale de la Santé (OMS). Weekly Epidemiological Record, 2022, vol. 97, 50 [full issue]. Weekly Epidemiological Record/Relevé épidémiologique hebdomadaire. 2022;97(50):645–74.

7. Markowitz LE, Gee J, Chesson H, Stokley S. Ten Years of Human Papillomavirus Vaccination in the United States. Academic Pediatrics. 2018;18(2):S3–S10. doi: 10.1016/j.acap.2017.09.014.

8. McLemore MR. Gardasil: Introducing the new human papillomavirus vaccine. Clin J Oncol Nurs. 2006;10(5):559–60. doi: 10.1188/06.Cjon.559-560. PubMed PMID: 17063609.

9. PATH. Projected and current national introductions, demonstration/pilot projects, gender-neutral vaccination programs, and global HPV vaccine introduction maps (2006-2023). 2022.

10. World Health Organization (WHO). HPV Dashboard. In: World Health Organization (WHO), editor. Immunization, Vaccines and Biologicals2025.

11. World Health Organization (WHO), UNICEF. Human Papillomavirus (HPV) vaccination coverage. In: World Health Organization (WHO), editor. Immunization data. https://immunizationdata.who.int/global/wiise-detail-page/human-papillomavirus-(hpv)-vaccination-coverage2025.

12. Meng X, Yang B, Yin H, Chen J, Ma W, Xu Z, et al. Global Burden and Incidence Trends in Cancers Associated with Human Papillomavirus Infection: A Population-Based Systematic Study. Pathogens. 2025;14(9). Epub 20250903. doi: 10.3390/pathogens14090880. PubMed PMID: 41011780; PubMed Central PMCID: PMCPMC12472360.

13. Wigle J, Coast E, Watson-Jones D. Human papillomavirus (HPV) vaccine implementation in low and middle-income countries (LMICs): health system experiences and prospects. Vaccine. 2013;31(37):3811–7.

14. Amponsah-Dacosta E, Kagina BM, Olivier J. Health systems constraints and facilitators of human papillomavirus immunization programmes in sub-Saharan Africa: a systematic review. Health policy and planning. 2020;35(6):701–17.

15. Bruni L AG, Serrano B, Mena M, Collado JJ, Gómez D, Muñoz J, Bosch FX, de Sanjosé S.,. Cameroon: Human Papillomavirus and Related Cancers, Fact Sheet 2023. 2023 10 March 2023. Report No.

16. Bruni L AG, Serrano B, Mena M, Collado JJ, Gómez D, Muñoz J, Bosch FX, de Sanjosé S.,. Human Papillomavirus and Related Diseases in Cameroon 2023 10 March 2023. Report No.

17. Ministry of Public Health Cameroon. DHIS-2 immunization portal. 2022.

18. Amani A, Nolna SK, Ndje MN, Ndongo CB, Ngounoue MD, Tiedeu B, et al. Social media controversy affecting the introduction of HPV vaccination for young girls in Cameroon. ARCH Women Health Care. 2019;10.

19. Ntonifor MM-N, Tazinkeng NN, Kemah B-L, Claudia NE, Sonia YK, Nchinjoh SC, et al. Factors associated with parental hesitancy towards the human papillomavirus vaccine: a cross-sectional study. Scientific Reports. 2025;15(1):18284. doi: 10.1038/s41598-025-94067-1.

20. World Health Organization (WHO)/Organisation mondiale de la Santé (OMS). Weekly Epidemiological Record, 2022, vol. 97, 24 [full issue]. Weekly Epidemiological Record/Relevé épidémiologique hebdomadaire. 2022;97(24):261–76.

21. Bruni L, Albero G, Rowley J, Alemany L, Arbyn M, Giuliano AR, et al. Global and regional estimates of genital human papillomavirus prevalence among men: a systematic review and meta-analysis. The Lancet Global Health. 2023;11(9):e1345–e62.

22. Njoh AA, Waheed D-EN, Kedakse TSNJ, Ebongue LJ, Kongnyuy EJ, Amani A, et al. Overcoming challenges and achieving high HPV vaccination uptake in Cameroon: lessons learned from a gender-neutral and single-dose program and community engagement. BMC Public Health. 2025;25(1):1696. doi: 10.1186/s12889-025-22776-3.

23. World Health Organization (WHO)/Organisation mondiale de la Santé (OMS). Guide to introducing HPV vaccine into national immunization programmes. Geneva2016.

24. Njoh AA, Saidu Y, Bachir HB, Ndoula ST, Mboke E, Nembot R, et al. Impact of periodic intensification of routine immunization within an armed conflict setting and COVID-19 outbreak in Cameroon in 2020. Confl Health. 2022;16(1):29. Epub 20220602. doi: 10.1186/s13031-022-00461-1. PubMed PMID: 35655226; PubMed Central PMCID: PMCPMC9161648.

25. Kelly SA, Changalucha J, Malibwa D, Ewing VL, Mkungu G, Deogratias D, et al. Knowledge and acceptability of male HPV vaccination among young people and community stakeholders in northwest Tanzania: social sciences in the Add-Vacc trial. Vaccine. 2026;69:128002. Epub 20251127. doi: 10.1016/j.vaccine.2025.128002. PubMed PMID: 41314053.

26. Iliassu S, Mbanga C, Ngenge MB, Ndoula S, Njoh AA, Griffith BC, et al. Stakeholders’ Perceptions of a Gender-Neutral Approach to Human Papillomavirus (HPV) Vaccination in Cameroon: A Qualitative Study 2025.

27. Pimenoff VN, Gray P, Louvanto K, Eriksson T, Lagheden C, Söderlund-Strand A, et al. Ecological diversity profiles of non-vaccine-targeted HPVs after gender-based community vaccination efforts. Cell Host & Microbe. 2023;31(11):1921–9.e3. doi: 10.1016/j.chom.2023.10.001.

28. Bogaards JA, Wallinga J, Brakenhoff RH, Meijer CJ, Berkhof J. Direct benefit of vaccinating boys along with girls against oncogenic human papillomavirus: bayesian evidence synthesis. Bmj. 2015;350:h2016. Epub 20150512. doi: 10.1136/bmj.h2016. PubMed PMID: 25985328; PubMed Central PMCID: PMCPMC4428278.

29. Xu MJ, Samuel O, Aslam N, and Van Loon K. Strategically striving to be more inclusive: A recommendation for gender-neutral human-papillomavirus vaccine policies. Human Vaccines & Immunotherapeutics. 2025;21(1):2480404. doi: 10.1080/21645515.2025.2480404.

30. Chesson HW, Ekwueme DU, Saraiya M, Dunne EF, Markowitz LE. The cost-effectiveness of male HPV vaccination in the United States. Vaccine. 2011;29(46):8443–50. Epub 20110802. doi: 10.1016/j.vaccine.2011.07.096. PubMed PMID: 21816193.

